# Background and concurrent factors predicting non-adherence to public health preventive measures during the chronic phase of the COVID-19 pandemic

**DOI:** 10.1101/2020.10.13.20211904

**Authors:** Yehuda Pollak, Rachel Shoham, Haym Dayan, Ortal Gabrieli Seri, Itai Berger

**Affiliations:** The Seymour Fox School of Education, The Hebrew University of Jerusalem, Jerusalem, Israel; Special Education Department, Talpiot College, Holon, Israel; The Paul Baerwald School of Social Work and Social Welfare, The Hebrew University of Jerusalem, Jerusalem, Israel

## Abstract

To determine factors that predict non-adherence to preventive measures for COVID-19 during the chronic phase of the pandemic, a cross-sectional, general population survey was conducted in Israel. Sociodemographic, health-related, behavioral, and COVID-19-related characteristics were collected. Among 2055 participants, non-adherence was associated with male gender, young age, bachelorhood, being employed, lower decrease in income, low physical activity, psychological distress, ADHD symptoms, past risk-taking and anti-social behavior, low pro-sociality, perceived social norms favoring non-adherence, low perceived risk of COVID-19, low perceived efficacy of the preventive measures, and high perceived costs of adherence to the preventive measures. There appears to be a need for setting out and communicating preventive measures to specifically targeted at-risk populations.

---

Novel Coronavirus 2019 (COVID-19) outbreak has an enormous impact on public health and global economy. In response to the growing pandemic, most states took preventive measures to limit the spread of cases through community transmission of COVID-19. These measures included isolation of infected and suspected patients, use of personal protective equipment (face masks, gloves etc.), personal hygiene, restrictions on gathering and traveling, social distancing, as well as mandatory quarantine and lockdown (1). Some of the preventive measures depend on how people cooperate, i.e. whether they adhere to the measures or not (2).

Despite the potentially harmful consequences for individuals and public health, non-adherence to the preventive measures (non-APM) for the COVID-19 pandemic, mainly at the acute phase, has been reported around the world (3-7). For designing effective public health policy, it is mandatory to identify the factors that predict non-APM. Recently, several cross-sectional surveys were conducted, in an effort to identify predictors of APM during the early phases of the pandemic. For instance, immediately after the detection of the first COVID-19 patient in Hong-Kong, higher levels of adoption of social-distancing measures were associated with being female, living in the geographic regions in Hong Kong that share the border with mainland China, perceiving oneself as having a good understanding of COVID-19, and being more anxious (7). In a survey conducted in Israel in April 2020, male gender, not having children, high levels of ADHD symptoms, smoking, past risk-taking behavior, and current psychological distress levels, all predicted non-APM. On the other hand, pro-sociality, understanding of the instructions, high perceived risk of COVID-19, and high perceived efficacy of the preventive measures predicted adherence (5).

As the COVID-19 pandemic continues, preventive measures become a constant part of our life. The objective of this study was to identify predictors of non-APM at the chronic phase of the pandemic. The literature suggests different conceptualizations of non-APM. Firstly, preventive measures are prescribed by health agencies and therefore can be considered medical instructions and healthy lifestyle. Secondly, non-APM might endanger the non-adherent and his/her vicinity, and consequently should be considered as a risk-taking behavior. Finally, preventive measures are often set as laws or regulations, implying that non-APM often means illicit behavior. Potential predictors of non-APM were chosen for this study based on the literatures regarding the risk factors for non-adherence to medical instructions (8), engagement in risk-taking behavior (9, 10), and engagement in anti-social behavior (11). These included four groups of variables: 1. Sociodemographic factors: age, gender, marital status, parenthood, ethnicity, religiousness, education, place of living, background migration, pre-outbreak and current level of income, pre-outbreak and current percent of position. 2. Health factors: healthy lifestyle (daily hours of sleep, smoking, physical activity), medical risk factors for COVID-19, subjective health, psychological distress, attention-deficit/hyperactivity disorder (ADHD) symptoms. 3. Behavioral and personality factors: past risk-taking behavior, past anti-social behavior, level of pro-sociality. 4. Perceptions regarding the COVID-19 and the preventive measures: the perceived risk of COVID-19, the perceived efficacy of the preventive measures, the perceived norms regarding APM, and the perceived cost of APM.

## Methods

This study was approved by the ethics committee of the Seymour Fox School of Education at the Hebrew University of Jerusalem. From May 13 to 23, 2020, a sample of 2055 online panel respondents (https://www.panel4all.co.il) representing most of the adult Israeli population was recruited.

For the primary outcome measure, non-APM, respondents were asked to rate the extent of which they adhered to each of the 13 preventive measures that were released by the Israeli Ministry of Health at the corresponding period (e.g. social distancing, personal hygiene, facemask). A five-point Likert scale was used: 1=‘*Not at all*’, 2=‘*Somewhat*’, 3=‘*Moderately*’, 4=‘*Strictly*’, and 5=‘Very strictly’. Individual mean response scores were calculated.

## Measures

The following scales were used to measure the independent variables:

1. Sociodemographic factors: Respondents completed a questionnaire consisting of items regarding age, gender, marital status, number of children, ethnicity, religious affiliation and level of observance, type of education, place of living (country region, and type of community), and background migration. In addition, respondent reported on pre-treatment level of income (much above average, above average, average, below average, much below average), level of decrease in income since the onset of the coronavirus outbreak (on a 1 = not at all to 5 = extreme decrease Likert scale), and pre-treatment and current percent of position.
2. Health factors: Respondents were asked to report on their average number of daily hours of sleep, frequency of engaging in intensive physical activity, and smoking habits. They also reported whether they are chronically treated or followed up for any of the following reasons (that are considered risk factors for COVID-19 (12)): heart disease, lung disease, liver disease, AIDS, cancer, organ transplantation, diabetes, dialysis, steroid treatment, and prophylactic antibiotic treatment. Subjective health was probed by a single-item self-rated health (SRH) Likert scale describing their own health impression, ranging from 1 = poor to 10= excellent. This scale was found to reflect individuals" perceptions of their physical health and psychological well-being (13). An adapted version of the Kessler Screening Scale for Psychological Distress (K6) was used to probe for non-specific psychological distress (14). For the purpose of the current study, only the first part of the scale was used, in which respondents rated on a five-level Likert scale (1 = ‘*All the time*’, 5 = ‘*None of the time*’) the level of six psychiatric common symptoms during the period of the COVID-19 crisis. The questionnaire is sensitive to high levels of mental distress (Kessler et al., 200, 2010(, and is used in the annual US National Health Interview Survey (15). The Hebrew version of the Adult ADHD Self-Report Scale (ASRSv1.1) (16, 17) was used to measure the level of attention deficit/hyperactivity disorder (ADHD) symptoms. A total score of ASRS was created by averaging the responses to all 18 items. For both the K6 and the ASRS, average scores were dichotomized at the clinical suggested cutoff (14, 16).
3. Behavioral and personality factors: The pro-social subscale of the young adult Strengths and Difficulties Questionnaire (SDQ) (18, 19) was used for measuring pro-sociality. Respondents rated the extent to which a series of six attributes described them during six months reference period on a three-level response scale (0 = ‘*not true*’, 1 = ‘*somewhat true*’, or 2 = ‘*certainly true*’). A short form of the Adult Risk-Taking Inventory (ARTI) (20, 21) was used to measure past engagement in risky behavior. The short form consists of 14 items probing for the frequency of engagement in relatively frequent activities (e.g., sunbathing without sunscreen, smoking marijuana) with respect to their frequency during the preceding year on a rating scale, ranging from 1 (Not at all) to 7 (On a daily basis). Previous work has shown that the ARTI has good reliability and validity. Past anti-social behavior was assessed using 15-item 4-point frequency scale, ranging from 1 = Not at all to 4 = More than 5 times, adapted from Cho et al. (22). For each of the three scales, the average continuous scores were converted to categorical scores by grouping values into four groups with quartiles as cutoff points.
4. COVID-19-related perception factors: Perceptions regarding the COVID-19 and the preventive measures were assessed using several five-point Likert scales: Perceived risk of COVID-19 was assessed by a nine-item self-report questionnaire that was designed for this study based on the risk perception literature (23). For example, “How likely are you to get COVID-19?”. In this sample, the scale had good internal consistency (Cronbach"s α = .80). Perceived efficacy of the prevent measure scale was measured by a self-report questionnaire designed for this study. The scale consists of five items was composed for measuring participants" perceived efficacy of the instructions. For example, “To what extent you think that adhering to the preventive measures will reduce the chances that you or your loved ones will get COVI-19?”. In this sample, the scale had good internal consistency (Cronbach’s α = .83). Another scale composed for this study, consisted of seven items probing for the perceived costs of APM, including the perceived cost of APM on different domains of wellbeing (e.g., economic, social, spiritual). For example, “To what extent adhering to the preventive measures will impair your interpersonal relationship?”. In this sample, the scale had good internal consistency (Cronbach’s α = .84). Perceived norms regarding APM were measured by four questions regarding the descriptive (i.e., the prevalence of non-APM) and the injunctive (i.e., the tolerance toward non-APM) norms of the family/friends and the community/workplace they are embedded in.

### Analytic approach

First, unadjusted logistic regression analyses were used to calculate the associations between each of the independent variables and the primary outcome. Next, four adjusted models were examined using backward stepwise logistic regressions with probability of 0.05 for entry and 0.1 for removal. Multicolinearity was examined through Spearman’s rank correlation analysis. In the first model, only the sociodemographic variables were included. In the second model, health-related variables were entered in a second block. Similarly, in the third and fourth models, the second block consisted of the behavioral and personality, and the COVID-19 related perceptions variables, respectively. P values were not corrected, a p-value < 0.05 was considered statistically significant.

## Results

### Sample characteristics

Table 1 summarizes the sociodemographic, health-related, behavioral, and COVID-19 perceptions related variables.

**Table 1:** Sociodemographic, health-related, behavioral and personality, and COVID-19 related perceptions characteristics of the sample

| **Characteristic** | **N (%)** |
| --- | --- |
| **Adherence to preventive measures** |  |
| Adherent | 1273 (61.9) |
| Non-adherent | 782 (38.1) |
| ***Sociodemographic*** |  |
| **Gender** |  |
| Women | 1128 (55.4) |
| Men | 909 (44.6) |
| **Age** |  |
| 18-24 | 309 (16.5) |
| 25-34 | 529 (28.3) |
| 35-44 | 377 (20.1) |
| 45-54 | 263 (14.1) |
| 55-64 | 266 (14.2) |
| =>65 | 127 (6.8) |
| **Ethnicity** |  |
| Jewish | 1834 (89.2) |
| Non-Jewish | 210 (10.2) |
| **Religiousness** |  |
| Non-religious | 1501 (73.7) |
| Religious | 535 (26.3) |
| **Marital status** |  |
| Single | 583 (28.6) |
| Married or in a relationship | 1302 (92.3) |
| Divorced, or widowed | 157 (7.7) |
| **Having children** |  |
| Yes | 1253 (68.3) |
| No | 582 (31.7) |
| **Higher Education** |  |
| Yes | 1054 (51.7) |
| No | 983 (48.3) |
| **Region/district** |  |
| North | 351 (17.1) |
| Haifa | 316 (15.4) |
| Center | 591 (28.8) |
| Tel Aviv | 205 (10.0) |
| Jerusalem | 254 (12.4) |
| West Bank | 60 (2.9) |
| South | 277 (13.5) |
| **Place of living** |  |
| Urban | 1637 (83.4) |
| Rural | 325 (16.6) |
| **Country of birth** |  |
| Israel | 1794 (87.3) |
| Not Israel | 260 (12.7) |
| **Pre-outbreak level of income** |  |
| Much above average | 78 (3.8) |
| Below average | 292 (14.3) |
| Average | 611 (30.0) |
| Above average | 498 (24.4) |
| Much below average | 561 (27.5) |
| **Pre-outbreak percent of position** |  |
| 0% | 326 (16.2) |
| 25% | 145 (7.2) |
| 50% | 207 (10.3) |
| 75% | 236 (11.7) |
| 100% | 1101 (54.6) |
| **Current percent of position** |  |
| 0% | 762 (38.3) |
| 25% | 157 (7.9) |
| 50% | 168 (8.5) |
| 75% | 196 (9.9) |
| 100% | 705 (35.5) |
| **Decrease in income (1-5 scale)** | Mean = 2.49<br>SD = 1.51 |
| **Health** |  |
| **Daily hours of sleep** | Mean = 8.79<br>SD = 1.37 |
| **Smoking** |  |
| No | 1820 (89.0) |
| Yes | 225 (11.0) |
| **Regular physical activity (number of days a week of 30 min vigorous-intensity activity)** |  |
| 0 | 515 (26.3) |
| 1-2 | 757 (38.6) |
| 3 or more | 688 (35.1) |
| **Risk factors for COVID-19** |  |
| No | 1755 (87.2) |
| Yes | 258 (12.8) |
| **Self-rated health (1-10 scale)** | Mean = 8.02<br>SD = 1.92 |
| **ADHD screener** |  |
| Below cutoff | 1729 (84.1) |
| Above cutoff | 326 (15.9) |
| **Psychological distress (K6)** |  |
| Below cutoff | 1513 (75.2) |
| Above cutoff | 500 (24.8) |
| **Behavior and Personality** |  |
| **Risk-taking behavior (ARTI)** |  |
| 1<sup>st</sup> quartile | 517 (25.2) |
| 2<sup>nd</sup> quartile | 446 (21.7) |
| 3<sup>rd</sup> quartile | 521 (25.4) |
| 4<sup>th</sup> quartile | 571 (27.8) |
| **Anti-social behavior (ASQ)** |  |
| 1<sup>st</sup> quartile | 476 (25.0) |
| 2<sup>nd</sup> quartile | 544 (28.6) |
| 3<sup>rd</sup> quartile | 395 (20.7) |
| 4<sup>th</sup> quartile | 490 (25.7) |
| **Pro-sociality (SDQ)** |  |
| 1<sup>st</sup> quartile | 527 (26.1) |
| 2<sup>nd</sup> quartile | 337 (16.7) |
| 3<sup>rd</sup> quartile | 461 (22.8) |
| 4<sup>th</sup> quartile | 696 (34.4) |
| ***COVID-19-related perceptions*** |  |
| **Descriptive and injunctive norms** |  |
| 1<sup>st</sup> quartile | 523 (26.0) |
| 2<sup>nd</sup> quartile | 498 (24.7) |
| 3<sup>rd</sup> quartile | 540 (26.8) |
| 4<sup>th</sup> quartile | 453 (22.5) |
| **Perceived risk of COVID-19** |  |
| 1<sup>st</sup> quartile | 598 (31.3) |
| 2<sup>nd</sup> quartile | 433 (22.7) |
| 3<sup>rd</sup> quartile | 448 (23.4) |
| 4<sup>th</sup> quartile | 432 (22.6) |
| **Perceived efficacy of the preventive measures** |  |
| 1<sup>st</sup> quartile | 470 (23.4) |
| 2<sup>nd</sup> quartile | 471 (23.4) |
| 3<sup>rd</sup> quartile | 568 (28.2) |
| 4<sup>th</sup> quartile | 502 (25.0) |
| **Perceived cost of adherence to preventive measures** |  |
| 1<sup>st</sup> quartile | 574 (28.5) |
| 2<sup>nd</sup> quartile | 450 (22.4) |
| 3<sup>rd</sup> quartile | 542 (26.9) |
| 4<sup>th</sup> quartile | 447 (22.2) |

### Rate of APM

Individual mean scores of APM were calculated. Based on the shape of the histogram (see Supplementary materials), a mean score < 4 was considered as non-adherence. A significant minority of the participants (38.1%) reported a mean level < 4 and was consequently defined as non-adherent.

### Predictors of non-APM

Spearman’s correlation coefficients were examined for all independent variables. All correlations were < 0.7. The correlations among age, marital status, and having children were in the 0.53-0.69 range.

Table 2 presents the unadjusted and adjusted regression analysis results for non-APM. The following variables were found to predict non-APM after adjustment for sociodemographic variables.

**Table 2:** Non-adherence to the public health preventive measures of the Ministry of Health for the COVID- 19 pandemic by a range of sociodemographic, health-related, risk-related, and instruction-related factors

|  | N (%) | Unadjusted OR | Adjusted OR |
| --- | --- | --- | --- |
| **Gender** |  |  |  |
| Women | 391 (43.0) | Ref | Ref |
| Men | 382 (33.9) | **1.47 (1.23-1.77)** | **1.40 (1.12-1.75)** |
| **Age** |  |  |  |
| 18-24 | 142 (46.0) | **3.65 (2.22-6.00)** | **2.80 (1.47 -5.33)** |
| 25-34 | 232 (43.9) | **3.35 (2.08-5.40)** | **3.21 (1.80-5.72)** |
| 35-44 | 142 (37.7) | **2.59 (1.59-4.24)** | **2.85 (1.59-5.11)** |
| 45-54 | 79 (30.0) | **1.84 (1.10-3.09)** | 1.74 (0.94-3.19) |
| 55-64 | 86 (32.3) | **2.05 (1.23-3.43)** | **2.25 (1.24-4.07)** |
| =>65 | 24 (18.9) | Ref | Ref |
| **Ethnicity** |  |  |  |
| Jewish | 702 (38.3) | Ref |  |
| Non-Jewish | 75 (35.7) | 0.90 (0.67-1.21) |  |
| **Religiousness** |  |  |  |
| Non-religious | 562 (37.4) | Ref |  |
| Religious | 213 (39.8) | 1.11 (0.90-1.35) |  |
| **Marital status** |  |  |  |
| Single | 278 (47.7) | **2.13 (1.46-3.11)** | **1.90 (1.11-3.25)** |
| Married or in a relationship | 454 (34.9) | 1.25 (0.87-1.80) | 1.22 (0.77-1.94) |
| Divorced, or widowed | 47 (29.9) | Ref | Ref |
| **Having children** |  |  |  |
| Yes | 417 (33.3) | Ref |  |
| No | 270 (46.4) | **1.74 (1.42-2.12)** |  |
| **Higher Education** |  |  |  |
| Yes | 386 (36.6) | Ref |  |
| No | 390 (39.7) | 1.14 (0.95-1.36) |  |
| **Region/district** |  |  |  |
| North | 143 (40.7) | Ref |  |
| Haifa | 116 (36.7) | 0.84 (0.62-1.15) |  |
| Center | 215 (36.4) | 0.83 (0.63-1.09) |  |
| Tel Aviv | 72 (35.1) | 0.79 (0.55-1.13) |  |
| Jerusalem | 110 (43.3) | 1.11 (0.80-1.54) |  |
| West Bank | 18 (30.0) | 0.62 (0.35-1.13) |  |
| South | 108 (39.0) | 0.93 (0.67-1.28) |  |
| **Place of living** |  |  |  |
| Urban | 623 (38.1) | Ref |  |
| Rural | 123 (37.8) | 0.94 (0.78-1.27) |  |
| **Country of birth** |  |  |  |
| Israel | 705 (39.3) | **1.54 (1.16-2.04)** |  |
| Not Israel | 77 (29.6) | Ref |  |
| **Pre-outbreak level of income** |  |  |  |
| Much below average | 219 (39.0) | Ref |  |
| Below average | 182 (36.5) | 0.90 (0.70-1.15) |  |
| Average | 227 (37.2) | 0.92 (0.73-1.17) |  |
| Above average | 118 (40.4) | 1.06 (0.79-1.41) |  |
| Much above average | 29 (37.2) | 0.92 (0.57-1.51) |  |
| **Pre-outbreak position percent** |  |  |  |
| 0% | 112 (34.4) | Ref |  |
| 25% | 55 (37.9) | 1.17 (0.78-1.75) |  |
| 50% | 83 (40.1) | 1.28 (0.89-1.83) |  |
| 75% | 93 (39.4) | 1.24 (0.88-1.76) |  |
| 100% | 426 (38.7) | 1.21 (0.93 -1.56) |  |
| **Current percent of position** |  |  |  |
| 0% | 254 (33.3) | Ref | Ref |
| 25% | 76 (48.4) | **1.88 (1.33-2.66)** | **2.14 (1.41-3.26)** |
| 50% | 72 (42.9) | **1.50 (1.07-2.11)** | **1.82 (1.18-2.79)** |
| 75% | 71 (36.2) | 1.14 (0.82-1.58) | 1.09 (0.72-1.65) |
| 100% | 281 (39.9) | **1.33 (1.07-1.64)** | 1.21 (0.90-1.62) |
| **Decrease in income (1-5 scale)** | 2028 | 0.95 (0.89-1.01) | **0.91 (0.84-0.99)** |
| **Daily hours of sleep** | 2010 | **0.92 (0.86-0.99)** |  |
| **Smoking** |  |  |  |
| No | 667 (36.6) | Ref |  |
| Yes | 105 (46.7) | **1.51 (1.15-2.00)** |  |
| **Regular physical activity (number of days a week of 30 min vigorous-intensity activity)** |  |  |  |
| 0 | 219 (42.5) | Ref | Ref |
| 1-2 | 282 (37.3) | 0.80 (0.64-1.01) | **0.71 (0.53-0.94)** |
| 3 or more | 237 (34.4) | **0.71 (0.56-0.90)** | **0.68 (0.50-0.92)** |
| **Risk factors for COVID-19** |  |  |  |
| No | 667 (38.0) | Ref |  |
| Yes | 92 (35.7) | 0.90 (0.69-1.19) |  |
| **Self-rated health (1-10 scale)** | 2052 | 0.98 (0.94-1.03) |  |
| **ADHD screener** |  |  |  |
| Below cutoff | 619 (35.8) | Ref | Ref |
| Above cutoff | 163 (50.0) | **1.79 (1.42-2.28)** | **1.46 (1.04-2.05)** |
| **Psychological distress (K6)** |  |  |  |
| Below cutoff | 534 (35.3) | **1.50 (1.22-1.84)** | **1.41 (1.04-1.91)** |
| Above cutoff | 225 (45.0) | Ref | Ref |
| **Risk-taking behavior (ARTI)** |  |  |  |
| 1<sup>st</sup> quartile | 138 (26.7) | Ref | Ref |
| 2<sup>nd</sup> quartile | 132 (29.6) | 1.16 (0.87-1.53) | 0.92 (0.63-1.32) |
| 3<sup>rd</sup> quartile | 202 (38.8) | **1.74 (1.34-2.26)** | 0.97 (0.67-1.40) |
| 4<sup>th</sup> quartile | 310 (54.3) | **3.26 (2.53-4.21)** | **1.70 (1.16-2.50)** |
| **Anti-social behavior (ASQ)** |  |  |  |
| 1<sup>st</sup> quartile | 101 (21.2) | Ref | Ref |
| 2<sup>nd</sup> quartile | 177 (32.5) | **1.79 (1.35-2.38)** | **1.55 (1.09-2.21)** |
| 3<sup>rd</sup> quartile | 153 (38.7) | **2.35 (1.74-3.17)** | **2.11 (1.44-3.09)** |
| 4<sup>th</sup> quartile | 285 (58.2) | **5.16 (3.89-6.86)** | **3.71 (2.53-5.45)** |
| **Pro-sociality (SDQ)** |  |  |  |
| 1<sup>st</sup> quartile | 258 (49.0) | Ref | Ref |
| 2<sup>nd</sup> quartile | 122 (36.2) | **0.59 (0.45-0.78)** | 0.74 (0.51-1.07) |
| 3<sup>rd</sup> quartile | 173 (37.5) | **0.63 (0.49-0.81)** | 0.72 (0.51-1.01) |
| 4<sup>th</sup> quartile | 215 (30.9) | **0.47 (0.37-0.59)** | **0.61 (0.45-0.84)** |
| **Perceived non-adherence norms** |  |  |  |
| 1<sup>st</sup> quartile | 104 (19.9) | Ref | Ref |
| 2<sup>nd</sup> quartile | 158 (31.7) | **1.87 (1.41-2.49)** | 1.13 (0.77-1.64) |
| 3<sup>rd</sup> quartile | 239 (44.3) | **3.20 (2.43-4.21)** | **1.77 (1.22-2.56)** |
| 4<sup>th</sup> quartile | 264 (58.3) | **5.63 (4.23-7.48)** | **3.02 (2.02-4.52)** |
| **Perceived risk of COVID-19** |  |  |  |
| 1<sup>st</sup> quartile | 317 (53.0) | Ref | Ref |
| 2<sup>nd</sup> quartile | 191 (44.1) | **0.70 (0.55-0.90)** | 0.74 (0.53-1.04) |
| 3<sup>rd</sup> quartile | 146 (32.6) | **0.43 (0.33-0.55)** | **0.49 (0.35-0.70)** |
| 4<sup>th</sup> quartile | 90 (20.8) | **0.23 (0.18-0.31)** | **0.31 (0.21-0.47)** |
| **Perceived efficacy of the preventive measures** |  |  |  |
| 1<sup>st</sup> quartile | 295 (62.8) | Ref | Ref |
| 2<sup>nd</sup> quartile | 208 (44.2) | **0.47 (0.36-0.61)** | 0.75 (0.53-1.06) |
| 3<sup>rd</sup> quartile | 169 (29.8) | **0.25 (0.19-0.33)** | **0.48 (0.34-0.70)** |
| 4<sup>th</sup> quartile | 91 (18.1) | **0.13 (0.10-0.18)** | **0.28 (0.18-0.42)** |
| **Perceived cost of adherence to preventive measures** |  |  |  |
| 1<sup>st</sup> quartile | 181 (31.5) | Ref | Ref |
| 2<sup>nd</sup> quartile | 176 (39.1) | **1.40 (1.08-1.81)** | 1.32 (0.93-1.89) |
| 3<sup>rd</sup> quartile | 225 (41.5) | **1.54 (1.21-1.97)** | 1.35 (0.95-1.92) |
| 4<sup>th</sup> quartile | 182 (40.7) | **1.49 (1.15-1.93)** | **1.98 (1.35-2.89)** |

*Note* Adjust OR represents the results of a 2-block backward stepwise logistic regressions (probability of 0.05 for entry and 0.1 for removal) with the sociodemographic variables in the first block and one of the other group of variables in the second block. Bold values represent confidence intervals that do not contain zero

Sociodemographic variables: The following variables predicted non-APM on adjusted analyses: male gender, younger age (<=64), and being single. Being employed predicted more non-APM; conversely, decrease in income predicted more adherence.

Health-related variables: Less regular physical activity and higher levels of ADHD symptoms and of psychological distress predicted non-APM.

Behavioral and personality factors: High levels of past risk-taking behavior and anti-social behavior, as well as low levels of pro-sociality, predicted non-APM.

COVID-19 perception variables: Non-APM was predicted by higher perceived non-adherence norms, lower perceived risk of COVID-19, lower perceived efficacy of the preventive measures, as well as higher perceived costs of adherence.

## Discussion

This study aimed to identify risk factors for non-APM during the chronic phase of the COVID-19 outbreak. Several factors were found to predict non-adherence. Sociodemographic factors included male gender, young age, bachelorhood, being employed, and smaller decrease in income. Health-related factors included physical activity, psychological distress, and ADHD symptoms. Behavioral and personality factors included history of risk-taking and anti-social behavior, and low pro-sociality. Finally, COVID-19 perception factors included perceived social norms favoring non-adherence, low perceived risk of COVID-19, lower perceived efficacy of the preventive measures, and higher perceived costs of adherence to the preventive measures. Notably, the greatest predictors in terms of OR were lower age, past anti-social behavior, low perceived risk of COVID-19, the efficacy of the preventive measures, and the norms of adhering to the preventive measures.

The variables that predicted non-APM at the chronic phase of the outbreak in Israel were similar to those that predicted non-APM during the first wave in Israel (5), suggesting that similar motivations drive the decision whether to adhere to preventive measures or not. Many of the non-APM predictors that were found in this study have also been reported by studies conducted in other states. For instance, male gender and young age were linked to non-adherence in the US, Somalia, Saudi Arabia, and Hong Kong during the COVID-19 outbreak (3, 4, 7, 24, 25). The negative correlation between employment and adherence in the current study parallels the findings of Porten et al. during the SARS outbreak in Germany (26). Several studies highlighted the association between adherence and perceptions about the infection and the preventive measures in a variety of states during the current pandemic (27-29). The negative correlation between adherence and the perceived costs of adherence resembles the findings of DiGiovanni et al. (30) reporting that perceived economic costs of the quarantine in Canada during the 2004 SARS outbreak were related to non-adherence. The role of social norms has been demonstrated in a study concerning quarantine in Senegal during Ebola outbreak (31) and in Australia during H1N1 outbreak (32). The negative correlation between adherence and past risk-taking behavior and unhealthy lifestyle is in line with a study reporting that among young adults with hazardous drinking, adherence to public policies is suboptimal (33). Our study adds new predictors of non-adherence including ADHD symptoms, general risk-taking behavior, previous engagement in crime, as well as low pro-sociality, which contributed for better prediction of non-APM.

Many of the above listed factors have been shown to predict non-adherence to medical treatment (8), risk-taking behavior (9, 10), and anti-social behavior (11). Accordingly, adherence to preventive measures may be analyzed in all the corresponding theoretical frameworks.

Notably, having medical risk factors for COVID-19 (i.e., background diseases) did not predict higher adherence to preventive measures. A similar independency between objective risk and adherence was found in a study reporting no effect of the total probable cases of SARS on likelihood of adherence (34).

### Public Health Implications

In deriving implication for public health, it is important to differentiate between predictors that preceded the COVID-19 outbreak, and therefore can be considered risk factors for non-APM, and other variables that coincided with the outbreak and hence their causal relations with non-APM cannot be determined based on a cross-sectional study. The latter include the economic consequence of COVID-19, as well as the perceptions regarding the pandemic and the preventive measures. Nevertheless, these coinciding predictors may still be used for targeting populations at-risk to non-APM.

The current findings of observable risk factors for non-APM suggest that the nature and the communication of the preventive measures should be targeted for different people. Policymakers may develop specific plans for populations at risk of non-adherence, focusing on messaging, fostering, and enforcing preventive measures, as well as on increased monitoring of infection rate.

Further research is warranted for identifying other risk factors for non-APM across longer periods and changing contexts and for examining the efficacy of public health policy in promoting APM.

## Data Availability

All data will be uploaded to an open repository and meanwhile is available from the corresponding author on request

## List of supporting information

Supplementary material is enclosed.

## Acknowledgments

This research was supported by the Ministry of Science & Technology, Israel.

## Supplementary materials

**Figure 1:**
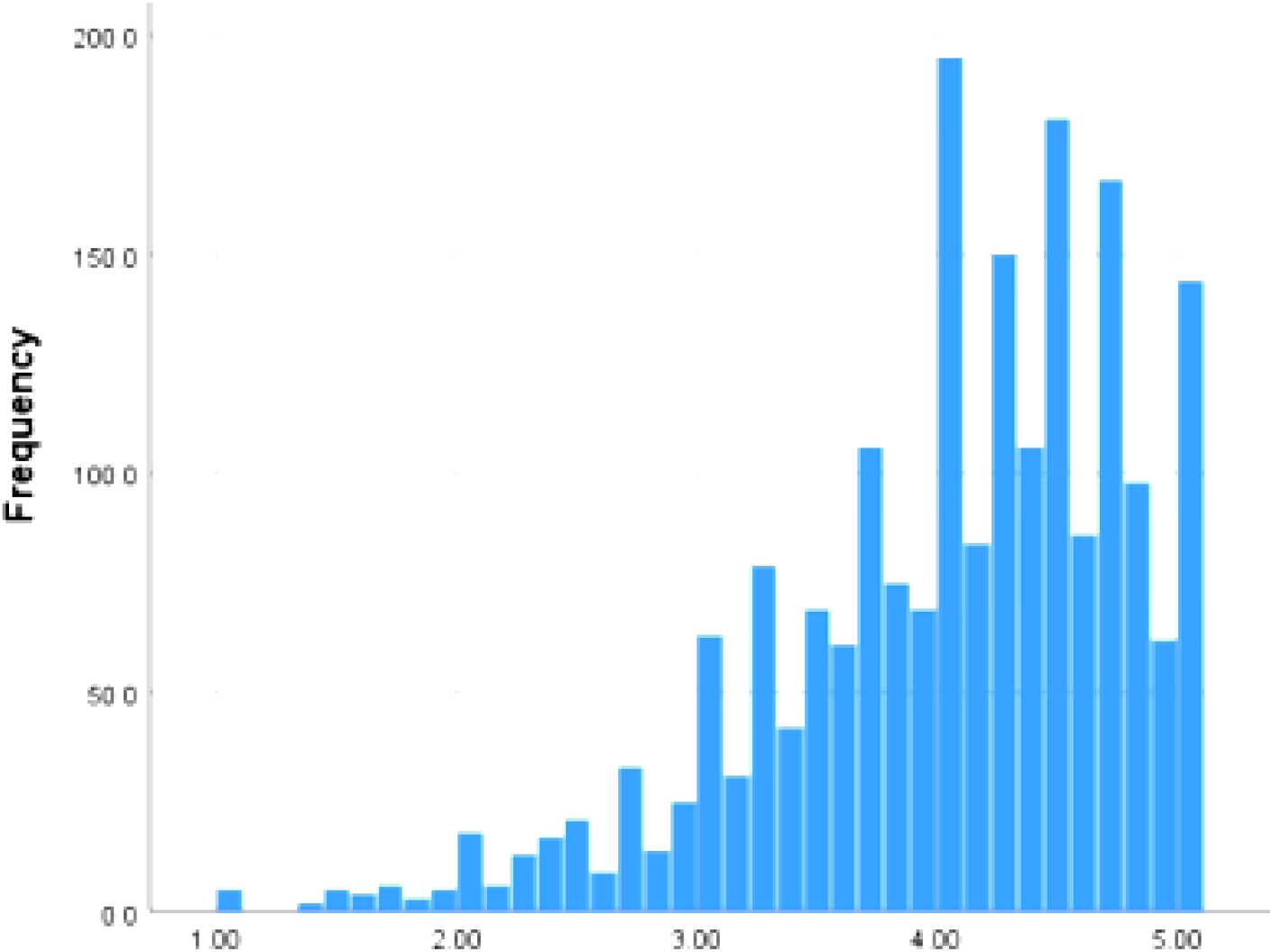
The distribution of the adherence to preventive measures scores (N = 2055)

## Notes

### Competing Interest Statement

The authors have declared no competing interest.

### Author Declarations

The ethics committee of the Seymour Fox School of Education at the Hebrew University of Jerusalem

## References

1. Pradhan D, Biswasroy P, Kumar Naik P, Ghosh G, Rath G. A Review of Current Interventions for COVID-19 Prevention. Arch Med Res. 2020 07;51(5):363–74.

2. Webster RK, Brooks SK, Smith LE, Woodland L, Wessely S, Rubin GJ. How to improve adherence with quarantine: rapid review of the evidence. Public Health. 2020 May;182:163–9.

3. Al-Hanawi MK, Angawi K, Alshareef N, Qattan AMN, Helmy HZ, Abudawood Y, et al. Knowledge, Attitude and Practice Toward COVID-19 Among the Public in the Kingdom of Saudi Arabia: A Cross-Sectional Study. Front Public Health. 2020;8:217.

4. Masters NB, Shih SF, Bukoff A, Akel KB, Kobayashi LC, Miller AL, et al. Social distancing in response to the novel coronavirus (COVID-19) in the United States. PLoS One. 2020;15(9):e0239025.

5. Pollak Y, Dayan H, Shoham R, Berger I. Predictors of non-adherence to public health instructions during the COVID-19 pandemic. Psychiatry Clin Neurosci. 2020 Jul.

6. Steens A, Freiesleben de Blasio B, Veneti L, Gimma A, Edmunds WJ, Van Zandvoort K, et al. Poor self-reported adherence to COVID-19-related quarantine/isolation requests, Norway, April to July 2020. Euro Surveill. 2020 09;25(37).

7. Kwok KO, Li KK, Chan HHH, Yi YY, Tang A, Wei WI, et al. Community Responses during Early Phase of COVID-19 Epidemic, Hong Kong. Emerg Infect Dis. 2020 07;26(7):1575–9.

8. Sabaté E, Sabaté E. Adherence to long-term therapies: evidence for action. World Health Organization; 2003.

9. Pollak Y, Dekkers TJ, Shoham R, Huizenga HM. Risk-Taking Behavior in Attention Deficit/Hyperactivity Disorder (ADHD): a Review of Potential Underlying Mechanisms and of Interventions. Curr Psychiatry Rep. 2019 Mar;21(5):33.

10. Figner B, Weber EU. Who takes risks when and why? Determinants of risk taking. Current Directions in Psychological Science. 2011;20:211–6.

11. Winters AM. Theoretical Foundations: Delinquency Risk Factors and Services Aimed at Reducing Ongoing Offending. Child and Adolescent Social Work Journal. 2020;37:263–9.

12. Jordan RE, Adab P, Cheng KK. Covid-19: risk factors for severe disease and death. BMJ. 2020 03;368:m1198.

13. Bjorner JB, Fayers P, Idler E. Self-rated health. In: Fayers PM, Hays RD, editors. Assessing Quality of Life in Clinical Trials: Methods and Practice. Oxford: Oxford University Press; 2005. p. 309–24.

14. Kessler RC, Andrews G, Colpe LJ, Hiripi E, Mroczek DK, Normand SL, et al. Short screening scales to monitor population prevalences and trends in non-specific psychological distress. Psychol Med. 2002 Aug;32(6):959–76.

15. Kessler RC, Green JG, Gruber MJ, Sampson NA, Bromet E, Cuitan M, et al. Screening for serious mental illness in the general population with the K6 screening scale: results from the WHO World Mental Health (WMH) survey initiative. Int J Methods Psychiatr Res. 2010 Jun;19 Suppl 1:4–22.

16. Zohar AH, Konfortes H. Diagnosing ADHD in Israeli adults: the psychometric properties of the adult ADHD Self Report Scale (ASRS) in Hebrew. Isr J Psychiatry Relat Sci. 2010;47(4):308–15.

17. Kessler RC, Adler L, Ames M, Demler O, Faraone S, Hiripi E, et al. The World Health Organization Adult ADHD Self-Report Scale (ASRS): a short screening scale for use in the general population. Psychol Med. 2005 Feb;35(2):245–56.

18. Brann P, Lethbridge MJ, Mildred H. The young adult Strengths and Difficulties Questionnaire (SDQ) in routine clinical practice. Psychiatry Res. 2018 06;264:340–5.

19. Goodman R, Scott S. Comparing the Strengths and Difficulties Questionnaire and the Child Behavior Checklist: is small beautiful? J Abnorm Child Psychol. 1999 Feb;27(1):17–24.

20. Shoham R, Sonuga-Barke E, Yaniv I, Pollak Y. ADHD Is Associated With a Widespread Pattern of Risky Behavior Across Activity Domains. J Atten Disord. 2019 Oct 4:1087054719875786.

21. Shoham R, Sonuga-Barke E, Yaniv I, Pollak Y. What Drives Risky Behavior in ADHD: Insensitivity to its Risk or Fascination with its Potential Benefits? J Atten Disord. 2020 Aug 27:1087054720950820.

22. Cho YI, Martin MJ, Conger RD, Widaman KF. Differential Item Functioning on Antisocial Behavior Scale Items for Adolescents and Young Adults from Single-Parent and Two-Parent Families. J Psychopathol Behav Assess. 2010 06;32(2):157–68.

23. Sjoberg L. Factors in risk perception. Risk Anal. 2000 Feb;20(1):1–11.

24. Qeadan F, Akofua Mensah N, Tingey B, Bern R, Rees T, Talboys S, et al. What Protective Health Measures Are Americans Taking in Response to COVID-19? Results from the COVID Impact Survey. Int J Environ Res Public Health. 2020 08;17(17).

25. Ahmed MAM, Siewe Fodjo JN, Gele AA, Farah AA, Osman S, Guled IA, et al. COVID-19 in Somalia: Adherence to Preventive Measures and Evolution of the Disease Burden. Pathogens. 2020 Sep;9(9).

26. Porten K, Faensen D, Krause G. SARS outbreak in Germany 2003: workload of local health departments and their compliance in quarantine measures--implications for outbreak modeling and surge capacity?J Public Health Manag Pract. 2006 2006 May-Jun; 12(3):242–7.

27. Chong YY, Chien WT, Cheng HY, Chow KM, Kassianos AP, Karekla M, et al. The Role of Illness Perceptions, Coping, and Self-Efficacy on Adherence to Precautionary Measures for COVID-19. Int J Environ Res Public Health. 2020 09;17(18).

28. Nguyen NPT, Hoang TD, Tran VT, Vu CT, Siewe Fodjo JN, Colebunders R, et al. Preventive behavior of Vietnamese people in response to the COVID-19 pandemic. PLoS One. 2020;15(9):e0238830.

29. Smith LE, Amlȏt R, Lambert H, Oliver I, Robin C, Yardley L, et al. Factors associated with adherence to self-isolation and lockdown measures in the UK: a cross-sectional survey. Public Health. 2020 Sep;187:41–52.

30. DiGiovanni C, Conley J, Chiu D, Zaborski J. Factors influencing compliance with quarantine in Toronto during the 2003 SARS outbreak. Biosecur Bioterror. 2004;2(4):265–72.

31. Desclaux A, Badji D, Ndione AG, Sow K. Accepted monitoring or endured quarantine? Ebola contacts’ perceptions in Senegal. Soc Sci Med. 2017 04;178:38–45.

32. Braunack-Mayer A, Tooher R, Collins JE, Street JM, Marshall H. Understanding the school community’s response to school closures during the H1N1 2009 influenza pandemic. BMC Public Health. 2013 Apr;13:344.

33. Suffoletto B, Ram N, Chung T. In-Person Contacts and Their Relationship With Alcohol Consumption Among Young Adults With Hazardous Drinking During a Pandemic. J Adolesc Health. 2020 Sep.

34. Hsu CC, Chen T, Chang M, Chang YK. Confidence in controlling a SARS outbreak: experiences of public health nurses in managing home quarantine measures in Taiwan. Am J Infect Control. 2006 May;34(4):176–81.

